# The Impact of the Affordable Care Act on Colorectal Cancer Incidence and Mortality: the case of Kaiser Permanente of Northern California

**DOI:** 10.1101/2020.10.26.20218503

**Authors:** Catherine Lee, Elizabeth H. Eldridge, Mary E. Reed, Jeffrey K. Lee, Lawrence H. Kushi, Donna Spiegelman

**Author notes:** Corresponding author: Catherine Lee, PhD, Division of Research, Kaiser Permanente Northern California, 2000 Broadway, Oakland, CA 94611.

## Abstract

**Background:** The Patient Protection and Affordable Care Act (ACA) eliminated cost sharing for preventive services, including colorectal cancer (CRC) screening for individuals aged 50 to 75 with private health insurance. The present study is the first to examine the impact of the no-cost CRC screening due to the ACA on CRC incidence and mortality.

**Methods:** We modeled trends in CRC incidence and CRC-related mortality in an open cohort of 2,113,283 Kaiser Permanente Northern California (KPNC) members aged 50 years and older between 2003 and 2016 using an interrupted time series design. Individual-level data were analyzed at the month-level. Analyses were adjusted for age, race/ethnicity and sex. As a sensitivity analysis, we considered a controlled approach, with a comparison group of KPNC members covered by health plans with pre-ACA zero cost-sharing for CRC screening.

**Results:** A total of 178,582,512 person-months were used in the analysis of CRC incidence, of which 48% occurred in the period before the ACA was passed into law (1/1/2003-3/31/2010) and 52% after (4/1/2010-12/31/2016). In primary analyses, the model for CRC incidence indicated a drop in the trend coinciding with the passage of the ACA (change in level incidence rate ratio, IRR: 0.83, 95% CI: 0.77-0.90, p-value < 0.0001), followed by a decrease in trend (change in slope IRR: 0.97/year, 95% CI: 0.93-1.00, p-value = 0.05). Results for CRC-related mortality were similar. Our controlled results indicate that free screening due to the ACA was associated with greater improvements in CRC outcomes among members previously covered by health plans with out-of-pocket costs for screening, compared to health plans with zero cost sharing for screening before the ACA went into effect.

**Conclusions:** We found that free CRC screening due to the ACA was associated with a decrease in age-, race/ethnicity- and sex-adjusted CRC incidence and CRC-related mortality, after accounting for contemporaneous competing interventions. Furthermore, these findings were robust to the addition of a comparison group with zero cost sharing both pre- and post-ACA.

## Introduction

Colorectal cancer (CRC) is the third most commonly diagnosed cancer and the second leading cause of cancer death in the US^1,2^. Screening can aid in preventing CRC and related mortality by detecting precancerous growths and early stage disease that can be effectively treated as shown in several large clinical trials that have demonstrated that screening by the fecal occult blood test and flexible sigmoidoscopy, can reduce colorectal incidence by 23% and mortality by up to 33%^3-5^. A systematic review by the US Preventive Services Task Force (USPSTF) found that most modalities of screening for CRC are associated with decreased CRC incidence and mortality^6^. Thus, the USPSTF recommends screening for colorectal cancer starting at age 50 years and continuing until age 75 years^7^.

Despite this recommendation, screening rates in the US over the past decade have remained relatively low, increasing from an estimated 65% in 2010 among US adults aged 50 to 75 years to 69% in 2018^8,9^. Although many factors may influence an individual’s decision to be screened for CRC^10,11^, high out-of-pocket costs have been identified as a major barrier to screening. The Patient Protection and Affordable Care Act (ACA), which was signed into law in 2010, mandated that CRC screening, among other evidence-based preventive services, be provided at no cost to those with private health insurance, thus potentially removing this barrier to screening.

Since the passage of the ACA, there have been a number of studies examining the impact of the elimination of cost sharing for CRC screening on receipt of screening^12-15^; a recent systematic review showed conflicting results on the impact of ACA on CRC screening rates^16^. However, there have been few studies focusing on the impact of the largest US health reform law since the inception of Medicare and Medicaid on colorectal cancer incidence or mortality. One study examined the impact of the ACA’s Medicaid expansion on CRC diagnoses and post-ACA survival, which showed a significant 6.7% increase in diagnoses and a 27% reduction in the risk of death in the period after the ACA was passed compared to the period before in individuals with Medicaid coverage^17^. Similarly, another study examined the differential effect of the ACA’s cost sharing reduction between those with private insurance and seniors covered by Medicare and found an 8% increase of early-stage CRC diagnoses among seniors^15^. In the present study, we aimed to examine the impact of the no-cost ACA CRC screening on both CRC incidence and mortality in the patient population of Kaiser Permanente of Northern California aged 50 and older between 2003 and 2016, using an interrupted time series design^18,19^. Should a beneficial impact be found, we planned, in addition, to perform a controlled analysis to strengthen the validity of the findings.

## Methods

### Study design

We used an interrupted time series design^18,19^ to examine CRC incidence and CRC-related mortality before and after the ACA was passed into law. The intervention of interest was the elimination of cost sharing for colorectal cancer screening due to the ACA.

### Data sources

Kaiser Permanente Northern California is an integrated health care delivery system with about 4.5 million members who are representative of the regional population^20^. For this study, we used an open cohort aesign including the experience of KPNC members who were 50 years or older between January 1, 2003 and December 31, 2016 with no prior CRC diagnosis. This study was approved by the KPNC Institutional Review Board.

### Outcomes

We identified members with a new diagnosis of colorectal cancer through the KPNC cancer registry, which reports to the Surveillance, Epidemiology and End Results program and captures >98% of cancers diagnosed among members compared with manual review. We obtained colorectal cancer-related mortality data from the KPNC mortality linkage file which includes data from multiple sources, including internal reporting, California state death records, and the Social Security Administration. Cause of death data were complete only through December 2015. Outcomes are defined in detail in Section A.1 of the Supplementary Appendix.

### Intervention

The intervention of interest was the elimination of cost sharing for CRC screening after the ACA was enacted. While the elimination of cost-sharing for CRC screening was not fully required under the ACA until after September 2010, the study setting preemptively removed cost-sharing after the passage of the ACA in March 2010, therefore our study uses an intervention start date of April 2010.

### Potential confounding interventions

During the study period, KPNC implemented a colorectal screening program in which members aged 50-75 years, who were due for screening, are mailed fecal immunochemical testing (FIT) kits annually at no cost ^21-24^. The program, which was rolled out in 2007, has been shown to be associated with reductions in both CRC incidence and CRC-related mortality^21^. To fully understand the impact of cost-free screening due to the ACA on CRC incidence and mortality in our population, we considered this program as an additional intervention potentially confounding the effects of the ACA.

### Statistical analysis

We used an interrupted time series model for our primary analyses of CRC incidence and mortality. We modeled both outcome rates separately, at the month level, using log-Poisson generalized linear models including time in month as a linear term and allowing for a change in the level and trend after both the KPNC FIT CRC screening program launch in January 2007 and the ACA launch in April 2010 through the inclusion of time-varying indicator variables and linear spline terms, respectively. We adjusted all models through covariate adjustment for categorical age (five-year age groups before age 85 and age 85+)^25^, race/ethnicity (Asian, Black, Hispanic, Indian, Multiracial, Pacific Islander, White, unknown), and sex (female, male, other/unknown). Details regarding model specification, presented effects and statistical comparisons can be found in Appendix Section B.1.

We standardized estimated monthly CRC incidence and mortality by direct standardization of model-based estimates to the distribution of person-months of age, race/ethnicity and sex over the cohort. We describe this methodology in detail in Appendix Section B.2. We report adjusted rates in terms of events per 100,000 person-years.

### Sensitivity analyses

We explored non-linear time trends using restricted cubic splines^26^. We accounted for possible impacts on CRC incidence and mortality due to new membership drawn from the ACA’s Health Insurance Marketplace and Medicaid expansion in two ways: 1) we allowed for a change in level and time trend at the beginning of 2014; and 2) we administratively censored our analyses immediately before 2014. Since the impact of intervention and competing interventions could have a lagged effect on outcome rate, we employed a data-driven model selection algorithm (described in detail in Appendix Section B.3) for each outcome, separately, that determined a best fitting model with: significant (p-value<0.05) segmented regression variables (change in level and/or slope attributed to the KPNC FIT CRC screening program and ACA); and an optimal lag of one to 12 months for each segmented regression variable included in the model.

We considered a controlled approach to account for possible underlying trends^18^. We hypothesized that there should be no effect of the ACA on colon cancer outcomes among members with no out-of-pocket costs for CRC screening before the ACA went into effect and defined the comparison group as member person-months tied to health plan coverage with zero cost sharing for screening before the ACA, specifically, no-deductible health plans with zero-dollar copayments. Conversely, we expected that the elimination of cost sharing for screening would have the greatest impact on those with prior out-of-pocket costs before the ACA. We stratified this group into two subgroups, the experience of members with high deductible insurance plans (≥$1000 yearly)^12^, who would have the most to gain from the elimination of cost sharing, and the experience of members with lower deductible plans (<$1000 yearly) or a copayment.

Due to incomplete insurance data prior to 2007, only data after January 1, 2007 could be used in the controlled analysis. Moreover, we found the high deductible stratum to be too small to study and omitted this stratum in the controlled analyses (details in Appendix Section A.2). We employed the analytical approaches described above separately within the comparison group ($0 copayments for CRC screening pre-ACA) and the remaining membership (lower deductible plans (<$1000 yearly) or a copayment pre-ACA), referred to as the intervention group.

## Results

A total of 178,582,512 person-months corresponding to 2,113,283 unique members were used in the analysis of CRC incidence, of which 48% occurred in the period before the ACA was passed into law (1/1/2003-3/31/2010) and 52% after (4/1/2010-12/31/2016). We present basic characteristics of the open cohort experience used in the analysis of CRC incidence in Table 1. In the post-ACA period, there was a shift in the racial/ethnic distribution with a slight increase in the number of person-months contributed by members of Asian and Hispanic race/ethnicity and corresponding decrease in Whites. The prevalence of person-months with a zero-dollar copayment increased from 8.9% before the ACA to 60.5% after the ACA. The prevalence of (any) deductible plans also increased from 3.9% before ACA to 15.7% in the period after. Basic characteristics of the open cohort experience used in the analysis of CRC-related mortality can be found in Appendix Table C.1.

**Table 1.** Basic characteristics of the CRC incidence cohort (person-months (p-m), stratified by the period before and after ACA was passed into law.

| Characteristic | Pre-ACA<br>1/1/2003-3/31/2010<br>N = 85,130,649 p-ms | Post-ACA<br>4/1/2010-12/31/2016<br>N = 93,451,863 p-ms |
| --- | --- | --- |
|  | % | % |
| Age, in years |  |  |
| 50-<55 | 20.0 | 16.8 |
| 55-<60 | 22.3 | 21.1 |
| 60-<65 | 17.4 | 18.9 |
| 65-<70 | 12.4 | 15.0 |
| 70-<75 | 10.0 | 10.3 |
| 75-<80 | 7.9 | 7.4 |
| 80-<85 | 5.7 | 5.4 |
| 85+ | 4.2 | 5.1 |
| Race/Ethnicity |  |  |
| Asian | 14.3 | 16.6 |
| Black | 7.4 | 7.3 |
| Pacific Islander | 0.3 | 0.4 |
| Hispanic | 9.5 | 11.5 |
| Indian | 1.0 | 0.9 |
| Multiracial | 0.1 | 0.1 |
| White | 67.5 | 63.4 |
| Sex |  |  |
| Female | 54.0 | 54.0 |
| Male | 46.0 | 46.0 |
| Other | 0.0 | 0.0 |
| Copayment |  |  |
| \$0 | 8.9 | 60.5 |
| Median (IQR), in dollars | 15 (10,100) | 0 (0,5) |
| Deductible |  |  |
| >\$0 | 3.9 | 15.7 |
| Median (IQR), in dollars | 1000 (500,1500) | 1000 (400,2000) |

### Primary analysis of CRC incidence and CRC-related mortality

The age-, race/ethnicity- and sex-adjusted monthly and model-based estimates of CRC incidence and mortality are displayed in Figures 1 and 2, respectively, accounting for both the KPNC FIT CRC screening program and the ACA coverage of screening. Estimated regression coefficients are presented in Appendix Table C.2. Prior to the KPNC FIT CRC screening program, adjusted CRC incidence was steady (slope IRR: 1.00/year, p-value = 0.73). Following the introduction of the KPNC FIT CRC screening program, adjusted incidence increased by 10% (95% CI: 1.01-1.21, p-value = 0.03) and remained stable (slope IRR: 1.00/year, 95% CI: 0.96-1.05, p-value = 0.88) until the ACA was passed into law, after which incidence dropped by 17% (95% CI: 0.77-0.90, p-value < 0.0001) and decreased 3% more each year (slope 95% CI: 0.93-1.00, p-value = 0.05). A likelihood ratio test comparing the fitted model to the one omitting both KPNC FIT CRC screening program variables (change in level, change in slope) suggests that accounting for the KPNC FIT CRC screening program does not significantly contribute information to the model (p-value = 0.08).

**Figure 1.**
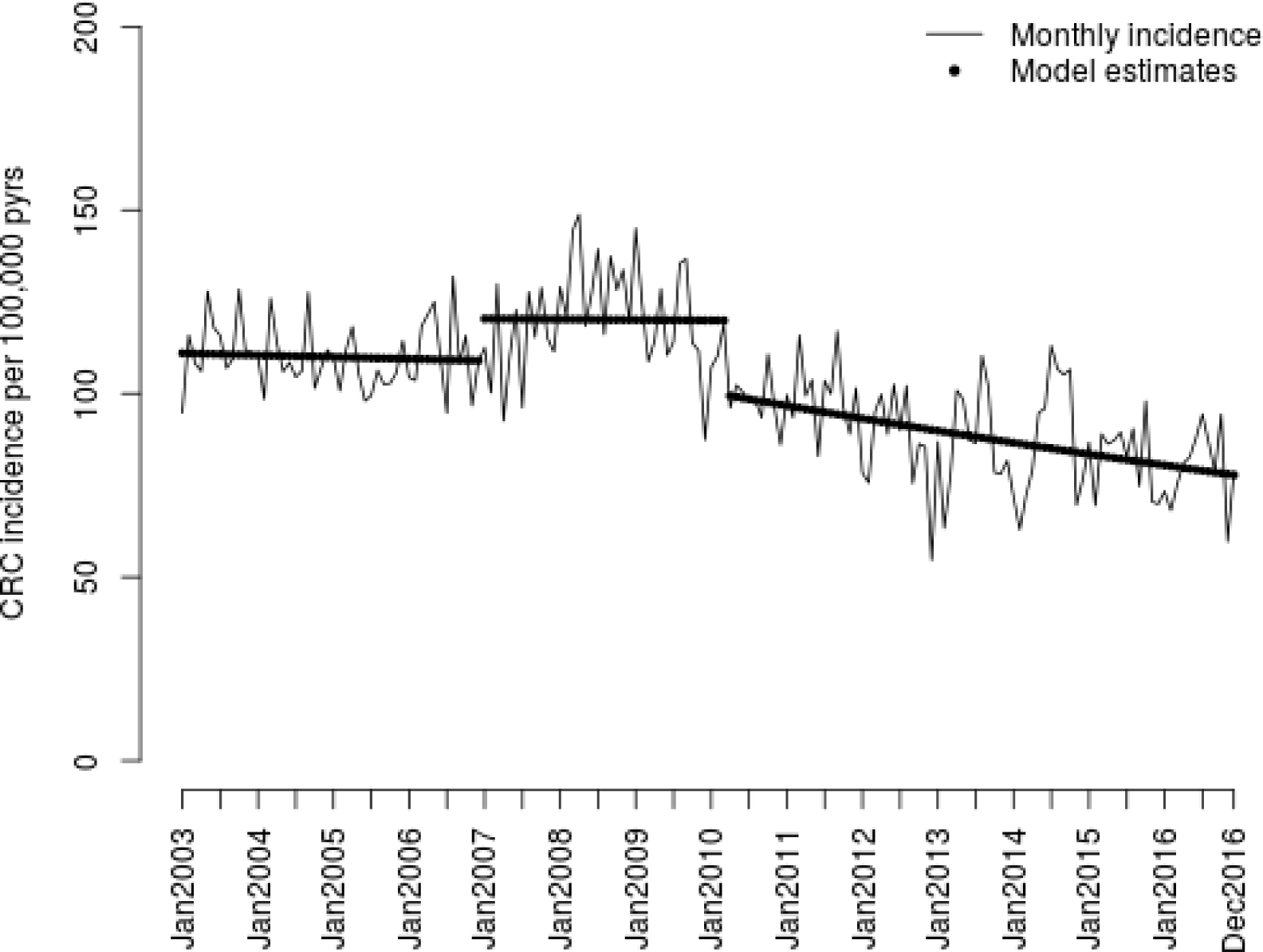
Age, race, and sex-adjusted monthly and model-based estimated CRC incidence in the entire cohort.

**Figure 2.**
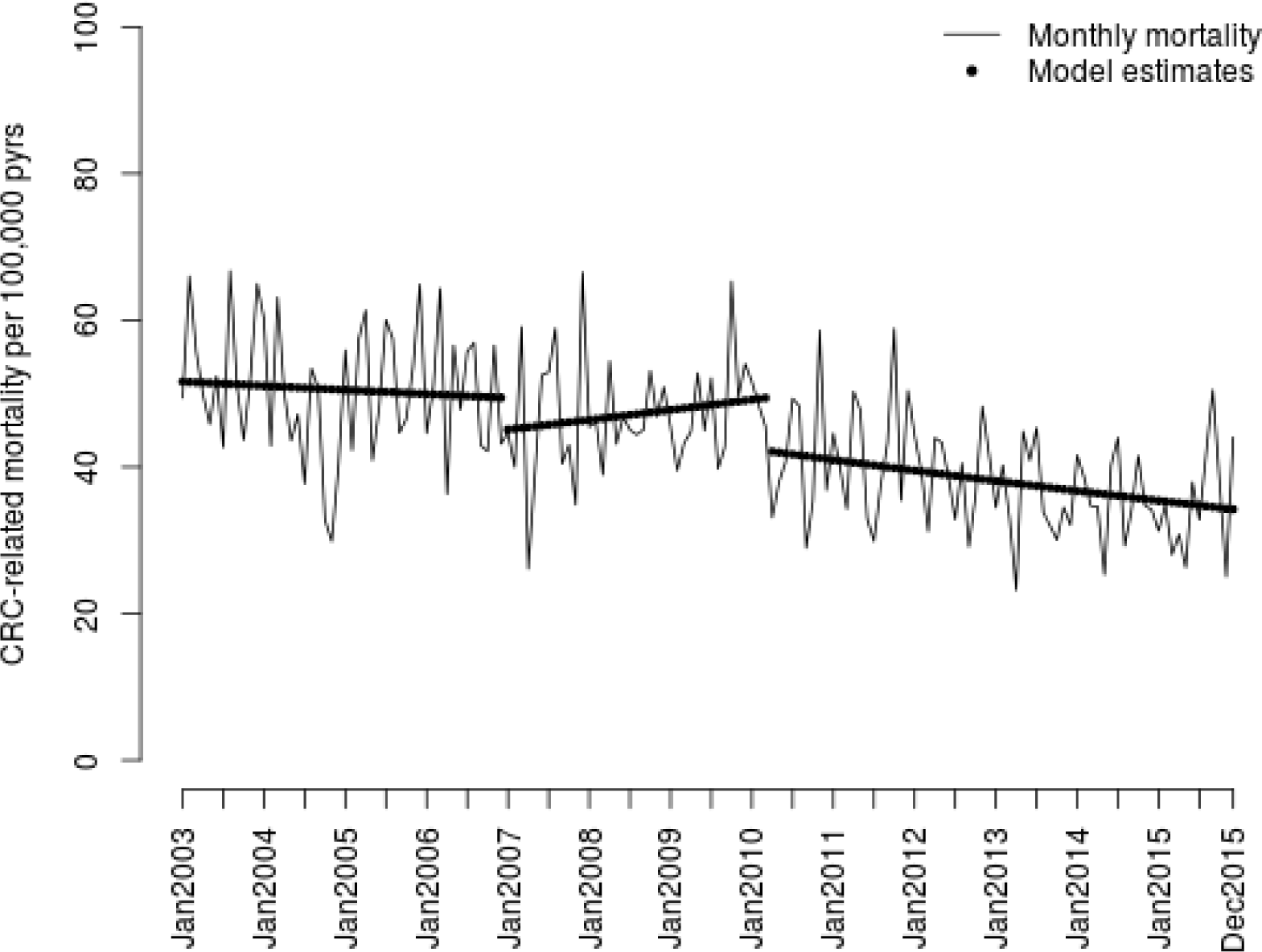
Age, race, and sex-adjusted monthly and model-based estimated CRC-related mortality in the entire cohort.

CRC-related mortality decreased over the study period, with our model estimating a drop in adjusted mortality upon launch of the KPNC FIT CRC screening program (level IRR: 0.91, 95% CI: 0.80-1.05, p-value = 0.19) followed by a slight increasing trend (slope IRR: 1.04/year, 95% CI: 0.97-1.11, p-value = 0.24). Following the introduction of the ACA, adjusted mortality decreased by 15% (95% CI: 0.75-0.96, p-value = 0.01) followed by a further 6% decreasing trend per year (95% CI: 0.88-0.99, p-value = 0.03). Similar to the analysis of incidence, a likelihood ratio test comparing the fitted mortality model to the one omitting both KPNC FIT CRC screening program variables (change in level, change in slope) suggests that accounting for the KPNC FIT CRC screening program does not significantly contribute information to the model (p-value = 0.26).

### Sensitivity analyses

Spline models were consistent with these main results. Findings were not impacted by accounting for the opening of the Health Insurance Marketplace and Medicaid expansion in the analyses (Appendix Table D.1). The model selection algorithm, which allowed for a lagged effect of the timing of significant intervention effects, indicated that our choice of timing of the impact of ACA was robust (Appendix Table D.2).

Basic characteristics of person-months by cost sharing strata (the zero-dollar copayments comparison group – for whom the ACA could have no impact; and the group with non-zero pre-ACA copayments – referred to as the intervention group) are presented in Appendix Table D.3. The direct-adjusted monthly and model-based estimates of CRC incidence and mortality in the comparison and intervention groups are displayed in Appendix Figures D.1 and D.2 with estimated effects in Table D.4. In both figures, it is apparent that the decrease in CRC incidence and mortality is greater in the intervention group compared to the zero-copayment comparison group, as hypothesized. We expressed the post-ACA trend as a slope, for ease of interpretation and comparison between stratified models. For incidence, within the comparison group stratum, the estimated trend in the post-ACA period was a 3% decrease in cases/year (95% CI: 0.95-0.99, p-value = 0.0009), a smaller effect than the trend in the stratum with prior out-of-pocket cost, a 7% reduction in cases/year (95% CI: 0.91-0.95, p-value < 0.0001). For mortality, within the comparison group, the trend in the post-ACA period was a 2% increase in CRC deaths/year (95% CI: 0.90-1.05, p-value = 0.18) compared to the trend observed in the intervention stratum, a 27% reduction in CRC deaths/year (95% CI: 0.69-0.77, p-value < 0.0001).

## Discussion

In a population-based cohort study of over 2 million members, we found that implementation of the ACA resulted in a significant drop in CRC incidence and related death by an estimated 22.5% and 23.5%, respectively (see Appendix E for details). Our findings remained consistent after accounting for potential influences in our sensitivity analyses. Secondary analyses also showed dramatic and significant reductions in CRC incidence and related death among those with no deductibles or co-payments before ACA implementation, further emphasizing the importance of reducing financial barriers to screening.

Our findings are consistent with a recent study examining the impact of the ACA’s Medicaid expansion on CRC incidence and overall survival in Kentucky, which showed a a significant 6.7% rise in CRC diagnoses,, which is expected when a new screening program is initiated and prevalence cases are detected. However, initiation of the ACA also resulted in a 27% reduction in post-diagnostic survival in the period after the ACA was passed compared to the period before among Medicaid recipients^17^. This harvesting effect may also explain the increase in diagnoses estimated in a recent study examining the impact of the ACA’s removal of cost-sharing on CRC diagnoses^15^. Our findings also align with previously published work from our team that examined age-adjusted outcome rates in a similar population before and after the KPNC organized CRC screening program and found decreases in incidence and mortality^21^. Note that in this prior work, mortality was examined only among members with a diagnosis (i.e., incidence-based mortality), differing importantly from the broader, population-based approach we took in this study. Our results complement these previous findings by incorporating the impact of both the ACA and the KPNC FIT screening program through an interrupted time series approach. Moreover, our controlled analysis provides insight into the outcome trends between the comparison and intervention groups during this time period, and further strengthens confidence in the results.

The changes in CRC incidence and mortality that we observed after implementation of the ACA and removal of cost-sharing for preventive services is likely due to increase uptake of screening. Although screening rates were rising prior to the ACA introduction, introduction of the ACA provided additional access to colonoscopies without the fear of a large co-payment or cost-sharing, which further lowered the risk of CRC and related death through early detection and removal of adenomatous polyps. In addition, the observed reduction in CRC incidence and mortality suggests a potentially substantial impact of this policy intervention. Based on the population attributable risk fraction^27^ due to the ACA (estimated at -22.5% for CRC-related mortality and -23.5% for incidence, see Appendix E for details), we estimate that the ACA may have prevented approximately 569 of 2,533 deaths from CRC in our health system population since its implementation. Should the benefits observed here apply to all of the US over this time period, approximately 65,327 of 290,346 deaths^25^ would have been prevented. In addition, the ACA would be associated with the prevention of extensive undue suffering and otherwise unnecessary medical procedures through the likely elimination of approximately 1,624 CRC cases among 6,912 diagnosed in KPNC and 177,102 of 753,627 in the US^25^ as a whole during the study period. Taken together, our results suggest the ACA has saved hundreds and thousands of lives and prevented much suffering.

Strengths of our study include that it was a population-based cohort study with a diverse study population and all screening encounters and cancer outcomes are captured with our electronic health records. In addition, our study had granular and detailed data on the cost-sharing, which allowed for improved understanding of the ACA’s impact. However, despite these strengths, we recognize that inferences from use of health care data should be made with caution. To emphasize, the data used in this study were not collected for research purposes; rather, we leveraged existing data sources available at KP collected for routine administrative purposes such as billing and internal disease and mortality registries. Other limitations to such include unmeasured confounding and analytic considerations, which are discussed in detail in Appendix F.

In a large, population-based cohort study, implementation of the ACA and reduction of cost-sharing was associated with a significant decrease in CRC incidence and related death. Our findings imply that removing barriers to screening, particularly financial burdens from cost-sharing, is an important health care policy that can result in improved CRC outcomes.

## Data Availability

The data used in this study contain Protected Health Information of Kaiser Permanente Northern California members and cannot be shared.

## Acknowledgements

The authors would like to thank Dr. Howard Koh for early discussions on this research topic that led to this paper, Jie Zhang for additional help with data management, Dr. Lawrence Gerstley for his help setting up a large data environment for analysis, and Dr. Theodore R. Levin and Dr. Douglas A. Corley for insight into the implementation of the KPNC organized FIT screening program. Lastly, the authors would like to thank Dr. Tracy Lieu, the director of the Division of Research, for her support.

