## Supplementary Appendix for "The Impact of the Affordable Care Act on Colorectal Cancer Incidence and Mortality: the case of Kaiser Permanente of Northern California"

Catherine Lee, PhD, Elizabeth H. Eldridge, MPH, Mary E. Reed, DrPH, Jeffrey K. Lee, MD, MPH, Lawrence H. Kushi, ScD, Donna Spiegelman, ScD

We present additional results that are beyond what could be presented in the main manuscript. This document is organized as follows:

Appendix A: Outcome-related information

| | | |
|---|---|---|
| Section A.1: | Detailed definition of outcomes | p. 1 |
| Section A.2: | Description of high deductible stratum | p. 1 |

Appendix B: Detailed description of statistical methods used in the manuscript

| | | |
|---|---|---|
| Section B.1: | The interrupted time series segmented regression model | p. 2 |
| Section B.2: | Direct adjusted outcome rates | p. 2 |
| Section B.3: | Data driven model selection algorithm to assess lagged effects and intervention effects | p. 2 |

Appendix C: Supplementary results

| | | |
|---|---|---|
| Table C.1: | Description of open cohort used in the CRC-related mortality analysis | p. 4 |
| Table C.2: | Interrupted time series regression coefficients for primary analyses of CRC incidence and CRC-related mortality | p. 5 |

Appendix D: Sensitivity analyses

| | | |
|---|---|---|
| Table D.1: | Regression coefficients for the analysis of CRC incidence and CRC-related mortality that: 1) allows for the impact of the Medicaid expansion and Heath Insurance Marketplace in 2014; and 2) administratively censors the data at the start of 2014 | p. 6 |
| Table D.2: | Regression coefficients from the analysis of CRC incidence and CRC-related mortality that allows for separate lags for the KPNC screening program and ACA | p. 7 |
| Table D.3: | Description of open cohort used in the analysis of CRC incidence, stratified by health insurance coverage | p. 8 |
| Table D.4: | Regression coefficients for the controlled analysis of CRC-incidence and CRC-related mortality | p. 9 |
| Figure D.1: | Age-, race/ethnicity- and sex-adjusted incidence from the controlled analysis of CRC incidence. | p. 10 |
| Figure D.2: | Age-, race/ethnicity- and sex-adjusted incidence from the controlled analysis of CRC-related mortality. | p. 11 |

Appendix E: Population attributable risk fraction calculations p. 12

Appendix F: Discussion regarding the study design and analytic approach p. 12

### Appendix A: Outcome-related information

#### Section A.1: Detailed definition of outcomes

Specifically, we used the International Classification of Diseases for Oncology (ICD-O-3) codes (C180, C181, C182, C183, C184, C185, C186, C187, C188, C189, C199, C209). We obtained mortality data from the KPNC mortality linkage file which includes data from multiple sources, including internal reporting, California state death records, and the Social Security Administration. We identified members who died of CRC-related causes based on ICD-9 (153.2, 153.3, 153.5, 153.6, 153.9) and ICD-10 codes (C18.0, C18.2, C18.3, C18.4, C18.5, C18.6, C18.7, C18.8, C19, C20). We defined death to be CRC-related if CRC was listed as any 'immediate', 'underlying', 'contributory', or 'other' cause of death.

#### Section A.2: Description of high deductible stratum

There were only 604 CRC diagnoses and 87 CRC-related deaths in the high deductible stratum over the entire study period. Although the crude incidence rate (64.2 per 100,000 p-yrs) and mortality rate (11.0 per 100,000 p-yrs) were comparable to publicly available data[1], an analysis within this stratum is likely underpowered.

#### Appendix B: Detailed description of statistical methods used in the manuscript

##### Section B.1: The interrupted time series segmented regression model

Individual-level data at the month-level were used in this analysis. We assumed a log-Poisson generalized linear model that allowed for a change in level and slope due to the ACA and the KPNC organized colorectal screening program:

$$\log(\text{cases}) = \log(\text{p-m}) + \beta_0 + \beta_1 \text{ month} + \beta_2 \, I(\text{month} \geq 1/2007) + \beta_3 \max(0, \text{month} - 1/2007) + \beta_4 \, I(\text{month} \geq 4/2010) + \beta_5 \max(0, \text{month} - 4/2010) + \beta \text{ confounders}$$

log(p-m): offset term which is the total person-months in a given month
$\beta_0$: baseline level
$\beta_1$: baseline slope
$\beta_2$: change in level after 1/1/2007, rollout of the KPNC organized CRC screening program
$\beta_3$: change in slope (relative to the baseline slope) after 1/1/2007
$\beta_4$: additional change (relative to the post KPNC organized CRC screening program trend line) in level after 4/1/2010, the passage of the ACA
$\beta_5$: additional change in slope (relative to the post KPNC organized CRC screening program trend line) after 4/1/2010

and I(month $\geq$ *C*) is an indicator function taking on value 1 if the month coincides or occurs later than *C*, and 0 otherwise.

We present regression estimates as incidence rate ratios (IRR) and corresponding 95% Wald-based confidence intervals (CI). Slope effects are presented as IRRs per one-year increase. Note that the change in slope terms are relative to the trend in the prior period; to transform a change in slope estimate to an absolute slope estimate with corresponding 95% CIs, we used the multivariate Delta Method. We used likelihood ratio tests to assess the goodness-of-fit of nested models.

##### Section B.2: Direct adjusted outcome rates

To obtain age-, race/ethnicity- and sex-adjusted estimates of outcomes, we first used the fitted model in Section A.1 to calculate model-based outcome rates in each of the age, race/ethnicity, and sex strata. We then multiplied each rate by the proportion of the corresponding age, race/ethnicity, and sex stratum in the entire incidence cohort and summed across strata.

##### Section B.3: Data driven model selection algorithm to assess lagged effects and intervention effects

We allowed for a lag in impact of intervention variables of one to 12 months defined as follows:

$$\log(\text{cases}) = \log(\text{p-m}) + \beta_0 + \beta_1 \text{ month} + \beta_2 \, I(\text{month} \geq 1/2007 + L_1) + \beta_3 \max(0, \text{month} - (1/2007 + L_1)) + \beta_4 \, I(\text{month} \geq 4/2010 + L_2) + \beta_5 \max(0, \text{month} - (4/2010 + L_2)) + \beta \text{ confounders},$$

over possible all possible sets of lags $\{(L_1, L_2): L_1 = 1,\ldots,12$ months and $L_2 = 1,\ldots,12$ months, and retained the log-likelihood value for each model.

In addition, we wanted to let the data inform which of the four intervention effects to include in the model: I(month $\geq$ 1/2007 + $L_1$), max(0, month – (1/2007 + $L_1$)), I(month $\geq$ 4/2010 + $L_2$), and max(0, month – (4/2010 + $L_2$)). We considered all $2^4$=16 possible models.

After fitting all 16 possible models, each with varying lags, we chose the model with the highest log-likelihood (best fit) and where all intervention effects were significant.

#### Appendix C: Supplementary results

Table C.1. Basic characteristics of the CRC mortality cohort (person-months (p-m) in terms of person-months (p-ms), stratified by the period before and after ACA was passed into law.

| | Pre-ACA 1/1/2003-3/31/2010 N = 85,190,787 p-m | Post-ACA 4/1/2010-12/31/2015 N = 78,315,421 p-m |
|---|---|---|
| Characteristic | % | % |
| Age, in years | | |
| 50-<55 | 19.9 | 17.0 |
| 55-<60 | 22.2 | 21.1 |
| 60-<65 | 17.4 | 18.9 |
| 65-<70 | 12.5 | 14.8 |
| 70-<75 | 10.0 | 10.2 |
| 75-<80 | 8.0 | 7.5 |
| 80-<85 | 5.7 | 5.4 |
| 85+ | 4.3 | 5.1 |
| Race/Ethnicity | | |
| Asian | 14.3 | 16.4 |
| Black | 7.4 | 7.3 |
| Pacific Islander | 0.3 | 0.4 |
| Hispanic | 9.5 | 11.2 |
| Indian | 1.0 | 0.9 |
| Multiracial | 0.1 | 0.1 |
| White | 67.5 | 63.8 |
| Sex | | |
| Female | 54.0 | 54.0 |
| Male | 46.0 | 46.0 |
| Other | 0.0 | 0.0 |
| Copayment | | |
| $0 | 8.9 | 59.5 |
| Median (IQR), in dollars | 15 (10,100) | 0 (0,10) |
| Deductible | | |
| >$0 | 3.9 | 15.0 |
| Median (IQR), in dollars | 1000 (500,1500) | 1100 (499,2000) |

Table C.2. Interrupted time series regression coefficients for primary analyses of CRC incidence and CRC-related mortality.

| | CRC incidence | | | CRC-related mortality | | |
|---|---|---|---|---|---|---|
| Variable | IRR | 95% CI | p-value | IRR | 95% CI | p-value |
| Baseline slope | 1.00 | (0.97,1.02) | 0.73 | 0.99 | (0.95,1.03) | 0.58 |
| Δ level after FIT[1] | 1.10 | (1.01,1.21) | 0.03 | 0.91 | (0.80,1.05) | 0.19 |
| Δ slope after FIT[1†] | 1.00 | (0.96,1.05) | 0.88 | 1.04 | (0.97,1.11) | 0.24 |
| Δ level after ACA | 0.83 | (0.77,0.90) | <0.0001 | 0.85 | (0.75,0.96) | 0.01 |
| Δ slope after ACA[†] | 0.97 | (0.93,1.00) | 0.05 | 0.94 | (0.88,0.99) | 0.03 |

The impact of the KPNC FIT CRC screening program was set to January 2007. The impact of the ACA was set to April 2010.

Δ stands for the phrase: "change in."

[1] "FIT" corresponds to the KPNC organized FIT CRC screening program.

[†] Change in slope effects are interpreted as the incidence rate ratio (IRR) corresponding to a one-year increase.

Δ level after ACA refers to the change in level of the trend line after KPNC FIT CRC screening program.

Δ slope after ACA refers to the change in level of the trend line after KPNC FIT CRC screening program. It is not relative to the baseline slope.

Incidence: Likelihood Ratio Test (LRT) comparing full model to the model excluding FIT terms corresponds to a p-value = 0.08.

Incidence: LRT comparing full model to the model excluding ACA terms corresponds to a p-value < 0.0001.

Mortality: LRT comparing full model to the model excluding FIT terms corresponds to a p-value = 0.26.

Mortality: LRT comparing full model to the model excluding ACA terms corresponds to a p-value = 0.02.

#### Appendix D: Sensitivity analyses

Table D.1. Regression coefficients for the analysis of CRC incidence and CRC-related mortality that allows for: 1) the impact of the Medicaid expansion and Heath Insurance Marketplace in 2014; and 2) censors the data at the start of 2014.

| **Allows for the impact of the Medicaid expansion and Health Insurance Marketplace in 2014** | | | | | | |
|---|---|---|---|---|---|---|
| | CRC incidence | | | CRC-related mortality | | |
| Variable | IRR | 95% CI | p-value | IRR | 95% CI | p-value |
| Baseline slope | 1.00 | (0.97,1.02) | 0.73 | 0.99 | (0.95,1.03) | 0.58 |
| Δ level after FIT[1] | 1.10 | (1.01,1.21) | 0.03 | 0.91 | (0.8,1.05) | 0.19 |
| Δ slope after FIT[1†] | 1.00 | (0.96,1.05) | 0.88 | 1.04 | (0.97,1.11) | 0.24 |
| Δ level after ACA | 0.85 | (0.78,0.93) | <0.0001 | 0.84 | (0.72,0.97) | 0.02 |
| Δ slope after ACA[†] | 0.95 | (0.91,0.99) | 0.02 | 0.96 | (0.88,1.05) | 0.36 |
| Δ level after Medicaid expansion | 1.05 | (0.96,1.16) | 0.27 | 0.89 | (0.76,1.04) | 0.13 |
| Δ slope after Medicaid expansion[†] | 1.00 | (0.96,1.07) | 0.58 | 1.01 | (0.92,1.11) | 0.78 |
| **Censors data at the start of 2014** | | | | | | |
| | CRC incidence | | | CRC-related mortality | | |
| Variable | IRR | 95% CI | p-value | IRR | 95% CI | p-value |
| Baseline slope | 0.99 | (0.97,1.02) | 0.71 | 0.99 | (0.95,1.03) | 0.60 |
| Δ level after FIT[1] | 1.10 | (1.01,1.21) | 0.03 | 0.91 | (0.8,1.05) | 0.19 |
| Δ slope after FIT[1†] | 1.00 | (0.96,1.05) | 0.84 | 1.04 | (0.97,1.11) | 0.24 |
| Δ level after ACA | 0.87 | (0.79,0.96) | 0.01 | 0.84 | (0.72,0.97) | 0.02 |
| Δ slope after ACA[†] | 0.92 | (0.87,0.98) | 0.01 | 0.96 | (0.88,1.05) | 0.35 |

The impact of the KPNC FIT CRC screening program was set to January 2007. The impact of the ACA was set to April 2010.
Δ stands for the phrase: "change in."
[1] "FIT" corresponds to the KPNC organized FIT CRC screening program.
[†] Change in slope effects are interpreted as the incidence rate ratio (IRR) corresponding to a one-year increase.
Δ level after ACA refers to the change in level of the trend line after KPNC FIT CRC screening program.
Δ slope after ACA refers to the change in level of the trend line after KPNC FIT CRC screening program. It is not relative to the baseline slope.

Table D.2 Regression coefficients for the analysis of CRC incidence and CRC-related mortality that allows for separate lags for the KPNC screening program and ACA.

| | CRC incidence* | | | CRC-related mortality⁑ | | |
|---|---|---|---|---|---|---|
| Variable | IRR | 95% CI | p-value | IRR | 95% CI | p-value |
| Baseline slope | 1.01 | (0.99,1.02) | 0.60 | 0.98 | (0.94,1.01) | 0.16 |
| Δ level after FIT[1] | 1.22 | (1.11,1.33) | <0.0001 | - | - | - |
| Δ slope after FIT[1†] | 0.90 | (0.85,0.96) | 0.03 | - | - | - |
| Δ level after ACA | 0.91 | (0.83,0.99) | <0.0001 | 1.42 | (1.28,1.57) | <0.0001 |
| Δ slope after ACA[†] | 1.06 | (1.00,1.13) | 0.04 | 0.88 | (0.85,0.91) | <0.0001 |

The impact of the KPNC FIT CRC screening program was set to January 2007. The impact of the ACA was set to April 2010.

Δ stands for the phrase: "change in."

[1] "FIT" corresponds to the KPNC organized FIT CRC screening program.

[†] Change in slope effects are interpreted as the incidence rate ratio (IRR) corresponding to a one-year increase.

Δ level after ACA refers to the change in level of the trend line after KPNC FIT CRC screening program.

Δ slope after ACA refers to the change in level of the trend line after KPNC FIT CRC screening program. It is not relative to the baseline slope.

[*] The data driven model selection process for CRC incidence selected changes in level and slope after FIT and ACA and selected the month of impact of the KPNC FIT program to be January 2008 (a one-year lag) and the month of impact of the ACA to be April 2010 (no lag).

⁑ The data driven model selection process for CRC-related mortality selected changes in level and slope after FIT and ACA and selected the month of impact of the KPNC FIT program to be January 2007 (no lag) and the month of impact of the ACA to be May 2010 (one-month lag).

Table D.3. Basic characteristics of the CRC incidence cohort (person-months (p-m), stratified by health insurance coverage ($0 copay and intervention) and pre-ACA (1/1/2003-3/31/2010) and post-ACA (4/1/2010-12/31/2016) periods.

| | Control group ($0 copayment) | | Intervention group (Copayment or deductible plan <$1000 annually) | |
|---|---|---|---|---|
| | Pre-ACA | Post-ACA | Pre-ACA | Post-ACA |
| Characteristic | % | % | % | % |
| Person-months | 6,735,885 | 44,246,620 | 76,420,865 | 39,796,882 |
| Age, in years | | | | |
| 50-<55 | 14.5 | 12.2 | 19.9 | 18.3 |
| 55-<60 | 17.0 | 14.5 | 22.4 | 25.4 |
| 60-<65 | 13.2 | 14.6 | 17.5 | 21.1 |
| 65-<70 | 15.5 | 18.9 | 12.4 | 13.2 |
| 70-<75 | 14.0 | 13.7 | 9.9 | 8.8 |
| 75-<80 | 11.2 | 10.4 | 7.9 | 5.9 |
| 80-<85 | 7.9 | 7.8 | 5.7 | 4.0 |
| 85+ | 6.6 | 7.9 | 4.2 | 3.2 |
| Race/Ethnicity | | | | |
| Asian | 16.4 | 15.1 | 14.0 | 17.9 |
| Black | 10.7 | 7.0 | 7.2 | 8.4 |
| Pacific Islander | 0.2 | 0.3 | 0.3 | 0.4 |
| Hispanic | 11.0 | 11.0 | 9.4 | 11.8 |
| Indian | 1.2 | 0.9 | 1.0 | 0.9 |
| Multiracial | 0.1 | 0.1 | 0.1 | 0.1 |
| White | 60.3 | 65.6 | 68.1 | 60.6 |
| Sex | | | | |
| Female | 58.3 | 55.6 | 53.8 | 53.1 |
| Male | 41.7 | 44.4 | 46.2 | 46.9 |
| Other | 0.0 | 0.0 | 0.0 | 0.0 |

Table D.4. Regression coefficients for the controlled analysis of CRC incidence and CRC-related mortality. The control arm consists of person-months corresponding to no-deductible plans with $0 copayments. The intervention arm consists of person-months corresponding to lower deductible plans (<$1000 yearly) or a copayment.

| **CRC incidence** | | | | | | |
|---|---|---|---|---|---|---|
| | Control ($0 copayment) | | | Intervention | | |
| Variable | IRR | 95% CI | p-value | IRR | 95% CI | p-value |
| Baseline slope | 1.04 | (0.94,1.14) | 0.46 | 0.99 | (0.96,1.03) | 0.73 |
| Δ level after ACA | 0.79 | (0.67,0.95) | 0.01 | 0.83 | (0.75,0.91) | 0.0001 |
| Δ slope after ACA[†] | 0.94 | (0.85,1.03) | 0.18 | 0.94 | (0.90,0.98) | 0.002 |
| **CRC-related mortality** | | | | | | |
| | Control ($0 copayment) | | | Intervention | | |
| Variable | IRR | 95% CI | p-value | IRR | 95% CI | p-value |
| Baseline slope | 1.05 | (0.91,1.20) | 0.52 | 1.02 | (0.97,1.09) | 0.44 |
| Δ level after ACA | 0.78 | (0.61,1.00) | 0.05 | 1.05 | (0.89,1.23) | 0.59 |
| Δ slope after ACA[†] | 0.98 | (0.85,1.12) | 0.73 | 0.71 | (0.65,0.77) | <0.0001 |

The impact of the ACA was set to April 2010.
Δ stands for the phrase: "change in."
[†] Change in slope effects are interpreted as the incidence rate ratio (IRR) corresponding to a one-year increase.

Figure D1. Age, race, and sex-adjusted monthly and model-based estimated CRC incidence in the control (person-months associated with $0 copay) and intervention groups (person-months associated with lower deductible plans (<$1000 yearly) or a copayment).

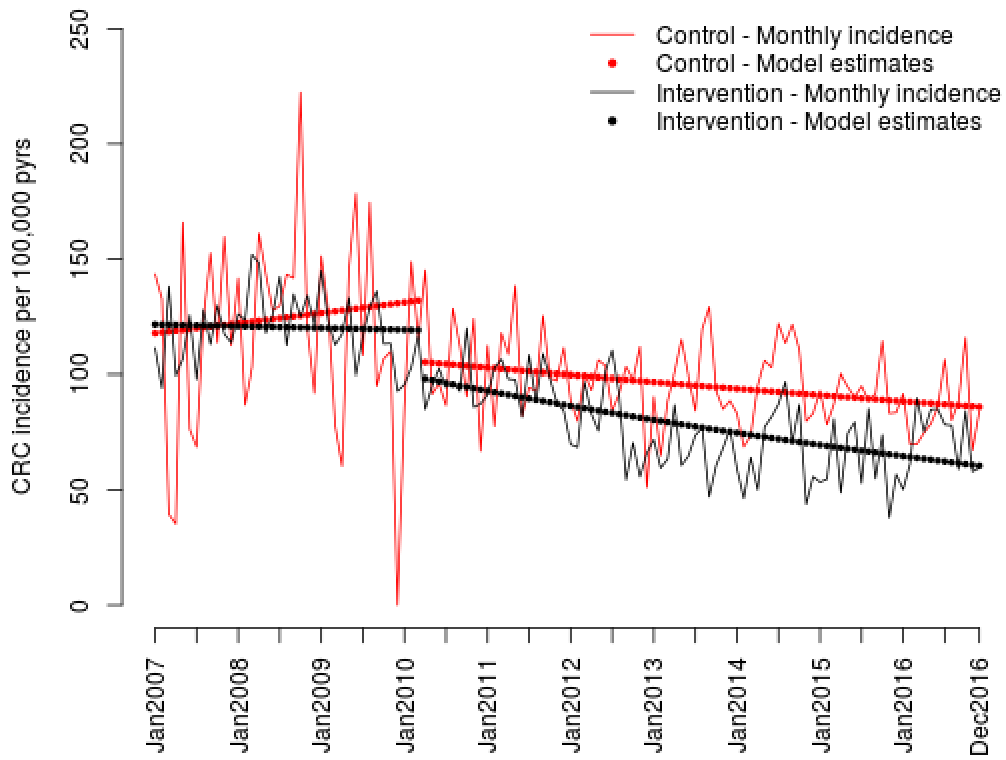

Figure D2. Age, race, and sex-adjusted monthly and model-based estimated CRC-related mortality in the control (person-months associated with $0 copay) and intervention groups (person-months associated with lower deductible plans (<$1000 yearly) or a copayment).

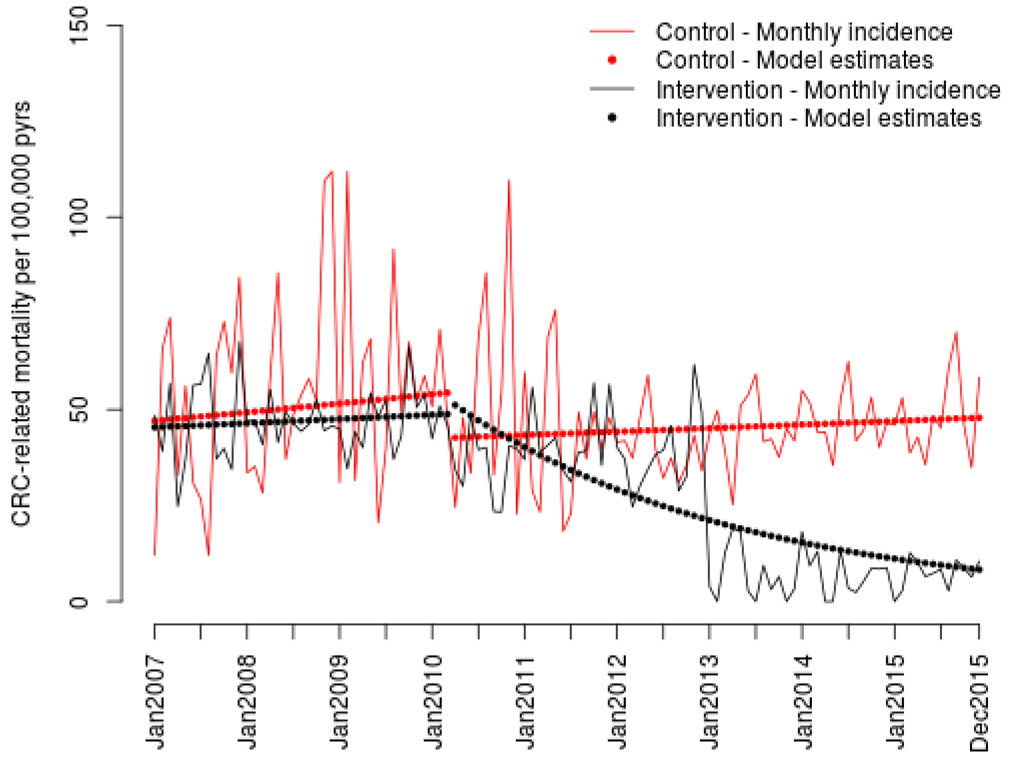

#### Appendix E: Population attributable risk fraction calculations

We directly standardized the CRC-related mortality and incidence rates before and after the ACA to the age-, race/ethnic-, and sex-distribution of the entire cohort. Direct adjusted mortality rates before and after ACA implementation were 48.9 per 100,000 p-yrs and 37.9 per 100,000 p-yrs, respectively. The associated population attributable risk percent (PAR)[2] due to the ACA can then be estimated at -22.5%. CRC incidence rates before and after ACA implementation were 115.0 per 100,000 person-years (p-yr) and 88.0 per 100,000 p-yrs, respectively, with the associated PAR due to the ACA of -23.5%.

#### Appendix F: Discussion regarding the study design and analytic approach

We had expected the prevalence of person-months in the post-ACA period with zero-dollar copayments to be close to 100% instead of the estimated 60.5% from our data (Table 1). This can be explained in part by grandfathered health plans that were in place before the ACA was signed into law, which were exempt from the ACA mandated removal of cost-sharing for CRC screening. While data on grandfathered health plan status were not readily available to us, the Kaiser Family Foundation 2019 Employer Health Benefits Survey[3] of private and non-federal public employers found that of all firms surveyed, 56% of covered workers were enrolled in grandfathered plans under the ACA in 2011 and 23% in 2016 (the end of our study period). In addition, regardless of grandfathered plan status, after the passage of ACA, copayments were largely not charged to members at the point of payment in the KPNC health care system; it is not possible to readily identify those for whom a copay may have still been in place post-ACA.

As these are health care data, our analyses may be subject to unmeasured confounding. Before-after designs, including interrupted time series, control for time-invariant confounding non-parametrically and are hence regarded as quasi-experimental[4]. In both controlled interrupted time series analyses of incidence and mortality, the intercept for the comparison and intervention groups roughly coincide (Figures C.1 and C.2), indicating that differences between these groups could be largely explained by age, race/ethnicity and sex. Note that it is not possible to conduct a randomized experiment of the ACA; these analytic approaches are thus the best available for assessing the causal impact of this policy intervention.

Although we find evidence of the impact on CRC incidence and mortality from elimination of cost-sharing for CRC screening by the ACA, our study was complicated by the presence of a competing intervention, the KPNC organized FIT CRC screening program. While we adjusted for this in our models and likelihood ratio tests indicated that including the KPNC organized screening program did not improve model fit, it is difficult to tease apart the contributions of both changes in policy that occurred contemporaneously, even if initial implementation differed by about three years. Nevertheless, our controlled analyses indicate that removing cost sharing for CRC screening was associated with substantially lower mortality among members with out-of-pocket costs for screening pre-ACA compared to members without, as defined by those with no-deductible plans with zero-dollar copayments.

Regarding our analytic approach, one might alternatively propose a difference-in-difference (DiD) approach comparing the change in outcome rate between members with $0 copayments for CRC screening in both pre- and post-ACA period (a comparison group unaffected by the removal of cost sharing for CRC screening due to the ACA) and members with out-of-pocket costs in the pre-ACA period and no out-of-pocket costs in the post-ACA period due to the ACA. But this design is not well-defined for our study, as we further explain. While our primary analysis is in terms of person-months, this ideal DiD analysis is in terms of individuals. When examining a short-term outcome such as screening[5], it is possible to observe outcomes from the same member in both pre- and post-periods and a difference in differences can be calculated. However, in the case of an event such as cancer incidence or mortality, these outcomes are only observed at most once per individual and must occur in either the pre- or post-period but not both, allowing only survivors into the post-period group. Furthermore, the definition of the two comparison groups presupposes that individuals did not experience the event in the pre-period, so that the same individuals can only contribute events to the post-period under this study design. Another approach for a controlled DiD analysis could be to replicate the study in another health care system where the ACA's removal of cost sharing for CRC screening were not present. However, this policy was applied to all states in the US in 2010 precluding such a comparator analysis. Another approach would be to replicate the analyses within the subset of members with grandfathered plans; as mentioned earlier in the discussion, these data were not available.

Nevertheless, we complemented our primary quasi-experimental pre-post analyses comparing monthly incidence and mortality rates in the pre- and post-ACA study periods with our best attempt at a control difference-in-difference analysis. The controlled analyses that we present were comparable to our primary uncontrolled analyses in that data were analyzed at the person-month-level, stratified by health coverage ($0 copayment vs. out-of-pocket costs as defined by lower deductible plans or a copayment). While a limitation to this approach is that the person-months with $0 copayments in the post-ACA period are a mixture of individuals who had $0 copayments in the pre-ACA period and out-of-pocket costs in the pre-ACA period, these findings were nevertheless consistent with the conclusions of the primary analyses.

The opening of the ACA's Health Insurance Marketplace and the Medicaid expansion in 2014 had the potential to bring in a new population of unscreened individuals into our membership who, in turn, could impact CRC outcome rates. However, we were not able to examine fully the effect of these programs on outcomes as we found that we had insufficient data to do so, particularly in the controlled analysis. While the controlled analysis of mortality (Figure C.2) suggests a decrease in mortality in the intervention group beginning in 2013, these findings should be interpreted with caution due to insufficient data after 2013.

Figure 6 in the American Cancer Society's (ACS) recent data[6] shows a sharply decreasing trend in CRC incidence and mortality between 1930 to the present. The ACS did not fit an interrupted time series analysis to the national data, as in our primary analysis, nor attempt to perform a controlled analysis, as in our secondary analysis. Future research could determine the extent to which, should these models be fit to the ACS data, that the same ACA effect as we have seen in KPNC, would be observed.

Finally, the observed improvements in CRC incidence and mortality in our analyses should have been mediated, at least in part, to increasing in screening, which we did not examine in this study. Potential future work would be to assess the extent to which screening mediates the impact of no-cost ACA preventive services on CRC incidence and mortality.

**Acknowledgements**

The authors would like to thank Dr. Howard Koh for early discussions on this research topic that led to this paper, Jie Zhang for additional help with data management, Dr. Lawrence Gerstley for his help setting up a large data environment for analysis, and Dr. Theodore R. Levin and Dr. Douglas A. Corley for insight into the implementation of the KPNC organized FIT screening program. Lastly, the authors would like to thank Dr. Tracy Lieu, the director of the Division of Research, for her support.

**Source of Funding:** This study was sponsored in part by NIH grant DP1 ES025459 (Dr. Donna Spiegelman, PI) and K07 CA212057 (Dr. Jeffrey Lee, PI).

**Disclosures**: No authors have any relevant disclosures.
